# Knowledge, Stigma and Prevalence of HBV among two populations from Nepal: a cross-sectional study

**DOI:** 10.1101/2023.08.13.23294044

**Authors:** Sam Hogan, Kate A McBride, Sameer Dixit, Andrew Page

## Abstract

**Background:** Hepatitis B virus (HBV) remains a public health issue in many developing nations, including Nepal. In Nepal the vaccination program was implemented in 2002-3 and aimed to reduce national prevalence of HBV. This study investigated socio-demographic, behavioural, and health service factors associated with HBV infection in low (Pokhara) and high (Dolpa) prevalence populations.

**Methods:** A serosurvey of 400 participants from within each population was conducted (final N = 799). The study combined a blood-draw to ascertain HBV status and a questionnaire that included demographic questions and items on knowledge of HBV, behavioural, and social factors associated with the risk of HBV infection. The blood draws were used to confirm HBV status as well as identify any previous infections from which participants may have recovered.

**Results:** There were 8/399 (2.01%, 95% CI 0.87%, 3.91%) current HBV infections among participants from Dolpa, compared with 2/400 (0.5%, 95% CI0.06%, 1.79%) among participants from Pokhara. The average age of each of the cohorts was relatively high, indicating an unclear association between vaccination programs and the low prevalence observed in this study. There was evidence from both sites there had been previous infections within the community as many of the participants indicated some level of protection from HBV either through vaccination or past infection (Dolpa 58/399, Pokhara 21/400) and recent cases who had recovered (Dolpa 4/399, Pokhara 3/400). Due to the relatively low prevalence of active cases of HBV, no meaningful associations between demographic, behavioural, and healthcare factors could be calculated. In both samples low level of knowledge of HBV and stigma towards people with HBV was evident.

**Conclusions:** This study found a low prevalence of HBV infection in both low and putatively high prevalence populations. However, knowledge of how HBV can be spread was quite low in each of the groups, indicating participants are managing to avoid infections while not necessarily being cautious around behaviours representing the greatest infection risk. There was also evidence of stigma being associated with having an active HBV infection, which may reduce the willingness of individuals to seek diagnosis or treatment.

## INTRODUCTION

Hepatitis B is a condition caused by the hepatitis B virus (HBV) when an individual becomes infected, and can become a chronic condition leading to severe complications. In 2015, the World Health Organisation (WHO) estimated that approximately 887,000 deaths globally were caused by HBV infections, with a further 257 million individuals continuing to live with chronic infections (1). The prevalence in South Asian contexts ranges from 1% to 6% (2).While these numbers vary due to geography and other external/demographic factors (3), several countries have vaccination programs in place to reduce new infections within their populations, including Nepal (4). The vaccines themselves are effective, however challenges exist in ensuring a high level of adherence to HBV vaccination programs to ensure ongoing population-level effectiveness, as multiple doses are required for the vaccine to be effective (5). Certain subgroups of the population are of increased importance to vaccination programs. The most common transmission pathway of HBV is from mother-to-child (1, 6), so it is of critical importance that children are vaccinated at an early age (7). There is some concern around how to incorporate those infants who have received a birth dose into the standard schedule of 6, 10 and 14 weeks within developing settings such as Nepal (8).

In the past, the national prevalence of HBV within Nepal has been estimated to be approximately 2-3% (4), however there have been sub-populations within Nepal which have recorded far higher prevalence levels than this. For example, in a previous study conducted in Dolpa, a remote region in Nepal, the prevalence of current HBV infection was estimated to be 18.7%, although the sample was based on those individuals who were able to travel to Kathmandu, and not the entire potential population of Dolpa (9, 10). While many risk factors for HBV are known (3), the reasons for the disparities between particular sub-populations and the general population of Nepal are less clear. Understanding the socio-demographic, knowledge, behavioural and health service factors that may explain these disparities is therefore important for ensuring vaccination program fidelity. Additionally, it has been shown there is a level of stigma attached HBV infections in other settings (11), even among those who have HBV (12). It is unclear whether this level of stigma exists within Nepal, so it is also important both knowledge of HBV and potential stigma towards those infected are assessed. Although the vaccination program is a national program run by the Family Welfare Division (FWD) in Nepal, many areas outside of major cities rely on community healthcare workers, especially those focussing on female health (Female Community Health Volunteers, FCHVs). These workers are deployed by the Ministry of Health and Population of Nepal (MOHP).

Accordingly, the current study sought to identify potential reasons for disparities in HBV prevalence in low and high prevalence populations and to ascertain the HBV prevalence in both areas to determine if the previously reported high prevalence in Dolpa is still reflective of the actual prevalence. This study also aimed to investigate socio-demographic and behavioural risk factors associated with HBV, as well as determine the relative knowledge of HBV among participants from a hypothesised high prevalence population (Dolpa) and low prevalence population (Pokhara).

## METHODS

### Study setting

Nepal has 77 districts in total. Dolpa is a remote and mainly undeveloped district of the Karnali Pradesh province in Mid-Western Nepal. Despite being the largest district in Nepal by land-size, the population in 2011 was only estimated to be 36,700, with much of the population living a semi-nomadic lifestyle. The degree to which the population of Dolpa differ from the rest of Nepal is unclear in terms of socio-cultural, lifestyle and infrastructure factors. Official statistics state that most of the population speak Nepali, however other sources state that there are very few Nepali speakers. This may in part be due to the semi-nomadic nature of the population, as migration across the border to Tibet and within Nepal is common. Dolpa was also chosen as it is an area which had a reported higher HBV prevalence previously (9).

Pokhara is a city in Nepal, located in the province of Gandaki Pradesh in Western Nepal. Pokhara has a population of approximately 403,000, and is a major hub for local and international tourism as it is located close to the Himalayas. Pokhara is located in a hills region of Nepal (elevation 1500m), while Dolpa is located in the mountains (70% of the district is between 3000m and 6400m). These sites are in provinces which border one another, however have vastly different geographic and socio-demographic features. Pokhara is a major urban area within Nepal and thus has far greater infrastructure than Dolpa. As a result, the lifestyles of the populations are drastically different, with the population of Pokhara being far more involved in industries such as tourism rather than self-sustaining agriculture.

### Participants

This study was a population-based, cross-sectional serological prevalence survey based on the Dolpa and Pokhara populations. Eligibility criteria for this study were being older 18 years old and living in either of the selected two districts. Sample size was calculated to be 400 participants from each site for a total of 800 participants. Participants were identified by with aid from local healthcare workers who helped recruit potential participants within their communities. In addition, local healthcare centers/clinics were used as the base for sampling and data collection. Of the 400 participants sampled at each site, only one participant did not complete the questionnaire. The final sample sizes were n = 399 from Dolpa and n = 400 from Pokhara. The study was implemented in accordance with ethical approval from the Western Sydney University Humar Research Ethics Committee (WSUHREC) (H13751) and the Nepal Human Research Council (NHRC) (Proposal ID 275 2021). Full demographics are listed in Table 1.

**Table 1.** A breakdown of the demographic details of the participant groups from Dolpa and Pokhara.

| Demographic info table |  |  |  |  |
| --- | --- | --- | --- | --- |
|  | Dolpa | % | Pokhara | % |
| Average age (years) | 35.35 | N/A | 47.19 | N/A |
| Literacy |  |  |  |  |
| Illiterate | 120 | 30.1 | 128 | 32 |
| Literate | 279 | 69.9 | 272 | 68 |
| Education level |  |  |  |  |
| No formal schooling | 121 | 30.3 | 125 | 31.4 |
| Primary school | 75 | 18.7 | 158 | 39.7 |
| High school | 183 | 45.9 | 102 | 25.6 |
| University | 20 | 5.1 | 13 | 3.3 |
| **Marital status** |  |  |  |  |
| Never married | 39 | 9.8 | 28 | 7 |
| Married | 337 | 84.4 | 337 | 84.3 |
| Divorced/Separated | 5 | 1.3 | 5 | 1.2 |
| Widowed | 18 | 4.5 | 30 | 7.5 |
| **Sex** |  |  |  |  |
| Male | 174 | 43.7 | 126 | 31.6 |
| Female | 224 | 56.3 | 273 | 68.4 |

### Questionnaire

A questionnaire was designed based on those used in the previous Integrated Biological Behavioural Survey (IBBS) series of studies (13, 14) which were performed by the Nepali Government to assess the prevalence of certain infectious diseases within at-risk subgroups among the general population. These questionnaires focus on demographic questions, as well as those which investigate drug use, alcohol consumption, sexual behaviours and knowledge of infectious diseases. The questionnaire used for this study also investigated many similar categories, including age, occupation, sexual behaviours and knowledge of the condition of interest. The section relating to knowledge and stigma associated with infectious diseases was instead modified, however, to focus solely on HBV.

These questionnaires were administered to participants in the field immediately after the blood draw had occurred, however there were some short wait times due to the high number of participants. Each participant was assigned a Participant ID (PID) number, which was used to link the blood draw to the questionnaire response. All questionnaires were conducted face-to-face by trained field staff from the Center of Molecular Dynamics Nepal (CMDN), the in-country research partner, and took approximately 15 minutes to complete. In return for taking part in the study, all participants were provided 100 Nepali Rupees, to re-imburse travel and work time. Participants were excluded if they were under the age of 18 or had already completed the questionnaire. The total number of field team members varied between sites, but always ranged between 4 and 6.

### Blood tests

To test for HBV cases, each participant provided a 5mL blood draw, which was taken by a trained phlebotomist. These blood samples were used in field test kits and also confirmed via laboratory test at the CMDN lab in Kathmandu. The rapid test kits used for initial diagnosis of HBV status were Qualisa ELISA HBsAg Test kits (15). These samples were transported via cold storage to ensure no degradation of the samples occurred between the field and the lab. The laboratory Enzyme-linked immunosorbent assays (ELISAs) were also able to identify different parts of the hepatitis B virus, namely the HBV surface (HBsAg) and core antigens (HBcAg), allowing some insight into the presence of active as well as previously recovered cases within the sample populations.

### Data Analysis

Descriptive statistics were used to investigate the demographic, behavioural and health service factors for each of the samples (Dolpa and Pokhara separately). These were used to measure features of the study sample to determine demographic and other information that might have affected the representativeness of the sample. Univariate and multivariate regression models were also used to investigate the association between each of the study factors on the likelihood of being infected with HBV. We also assessed the relationship between these variables and the recorded presence of HBsAb within the participants, as this indicates protection either from receiving a vaccine or through previous infection.

## RESULTS

### HBV Prevalence

In the Dolpa sample, 8/399 participants were HBV positive (2.01%, 95% CI 0.87%, 3.91%), in comparison to the Pokhara cohort which only had 2/400 participants as HBV positive (0.5%, 95% CI 0.06%, 1.79%). Overall, the Pokhara cohort was much older than the Dolpa cohort (average age of 47 vs 35), with a higher proportion of females (68.4% vs 56.3%). Both samples had the same number of married participants (337), but varied in some of the other relationship categories (Table 1). The distribution of education status was notable, with Dolpa having a higher proportion of participants having completed high school (45.9% vs 25.6%), but the percentage of participants who had no formal schooling was very similar between cohorts (30.3% for Dolpa vs 31.4% for Pokhara). The proportion of participants who self-identified as literate was also very similar between cohorts, with 69.9% of the Dolpa participants being literate compared to 68% from Pokhara. The prevalence of HBsAg within the two groups was 58/399 (14.5%, 95% CI 11.2%, 18.4%) in Dolpa and 21/400 (5.25%, 95% CI 3.28%, 7.91%). This indicates the number of participants who have some level of protection from HBV either from vaccination or being previously infected. The full results of these tests are available in Supplementary Table 1.

Due to the low number of positive cases, none of the univariate and multivariate regression models produced statistically significant results. Many of the models produced results which were entirely unrealistic, and are represented by the “N/A” entries in the tables. The results of these regressions are shown in Table 2.

**Table 2.**
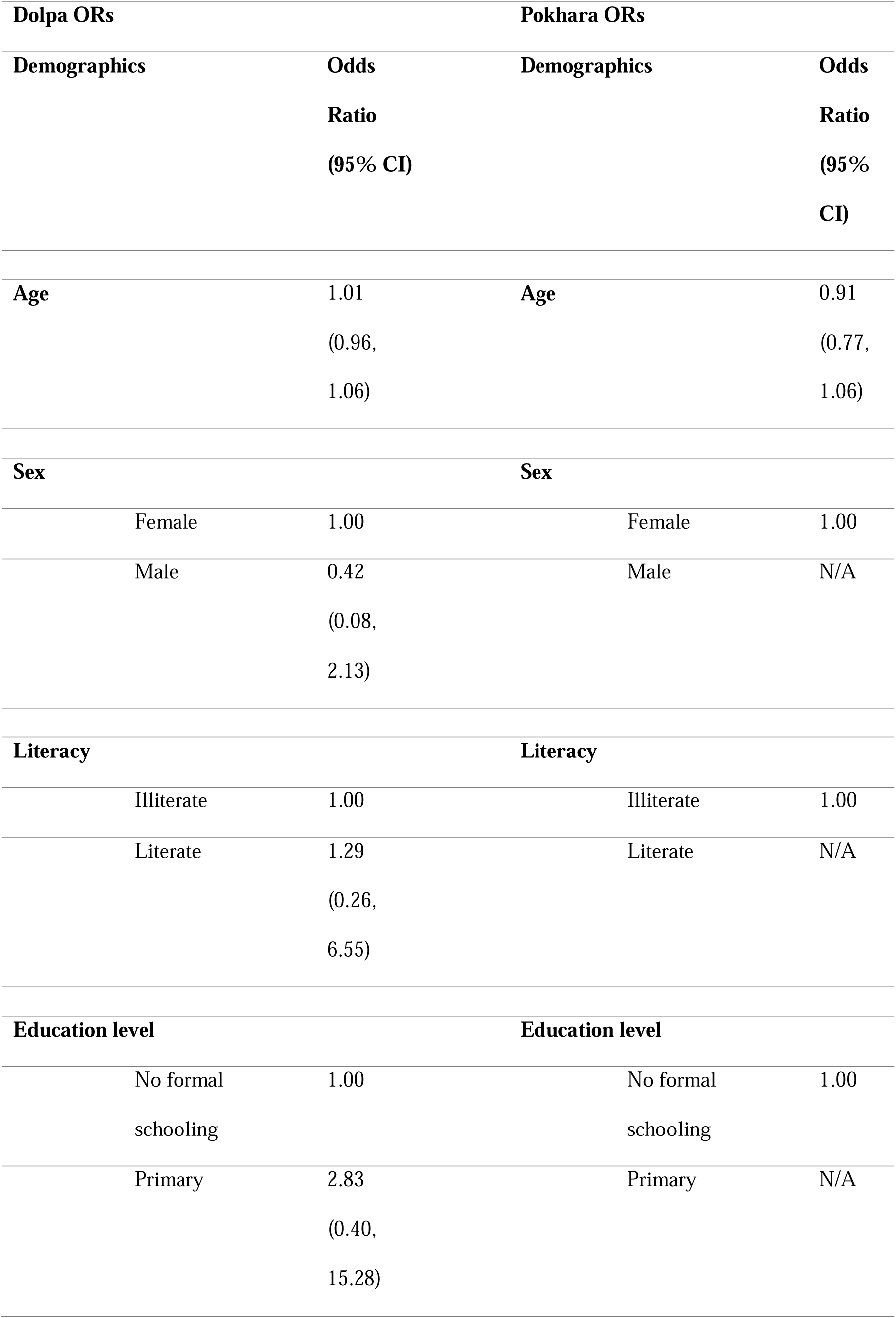
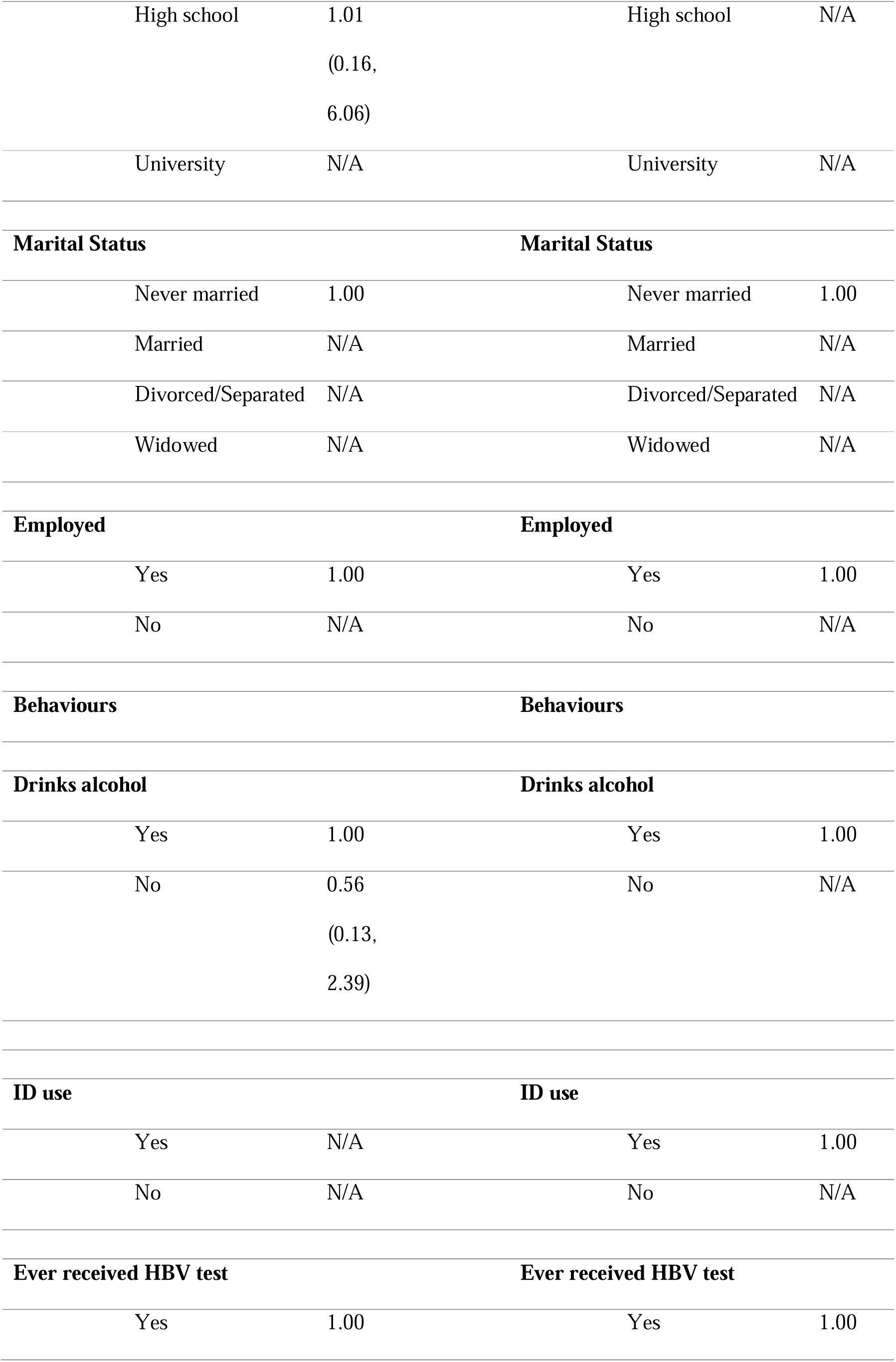
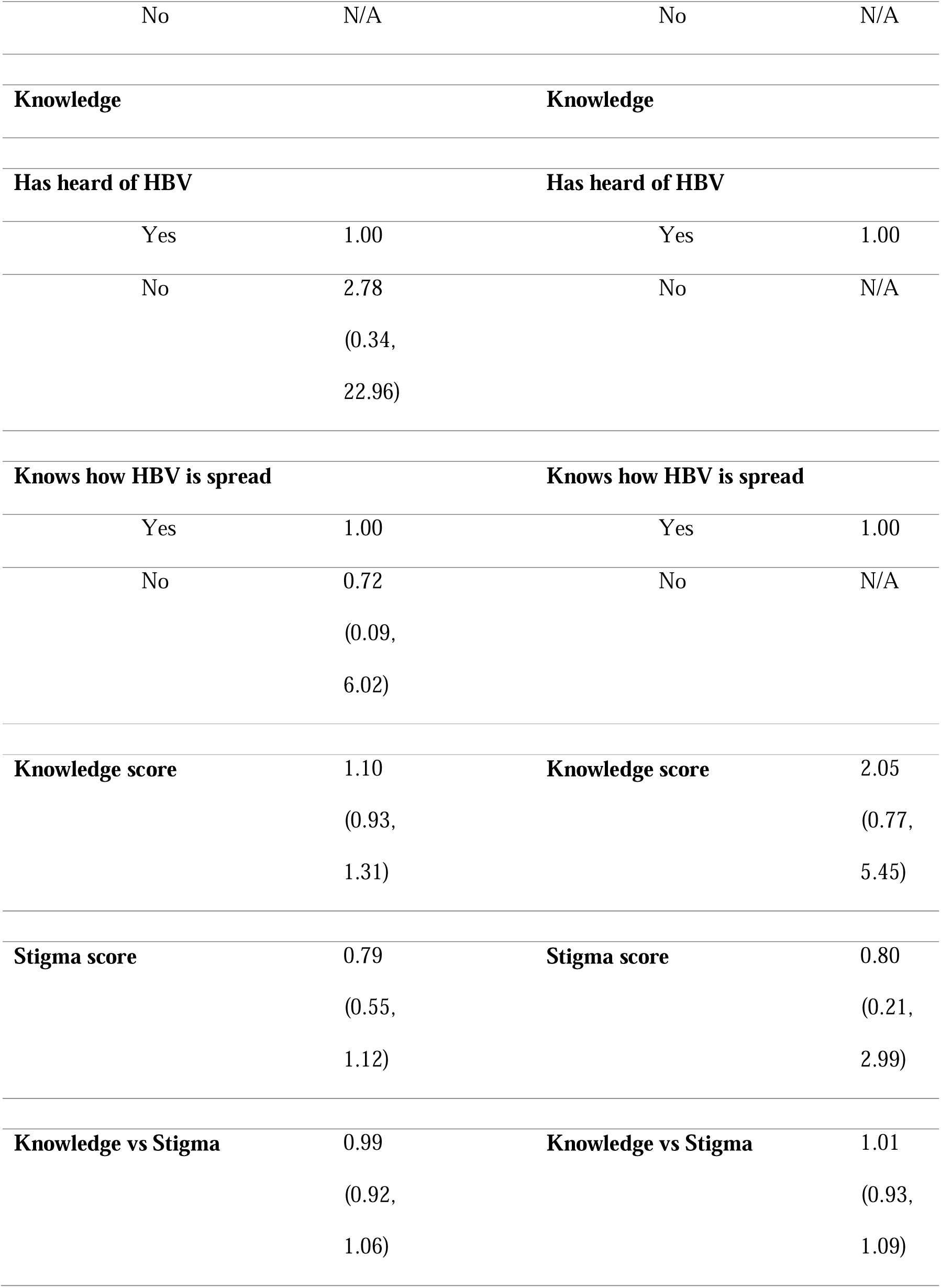
Regression models were used to produce odds ratios to indicate the increased or decreased likelihood of being infected with HBV based on different demographic, behavioural and knowledge-based risk factors.

| Dolpa ORs |  | Pokhara ORs |  |
| --- | --- | --- | --- |
| Demographics | Odds Ratio<br>(95% CI) | Demographics | Odds Ratio<br>(95% CI) |
| Age | 1.01<br>(0.96, 1.06) | Age | 0.91<br>(0.77, 1.06) |
| Sex |  | Sex |  |
| Female | 1.00 | Female | 1.00 |
| Male | 0.42<br>(0.08, 2.13) | Male | N/A |
| Literacy |  | Literacy |  |
| Illiterate | 1.00 | Illiterate | 1.00 |
| Literate | 1.29<br>(0.26, 6.55) | Literate | N/A |
| Education level |  | Education level |  |
| No formal schooling | 1.00 | No formal schooling | 1.00 |
| Primary | 2.83<br>(0.40, 15.28) | Primary | N/A |
| High school | 1.01<br>(0.16,<br>6.06) | High school | N/A |
| University | N/A | University | N/A |
| **Marital Status** |  | **Marital Status** |  |
| Never married | 1.00 | Never married | 1.00 |
| Married | N/A | Married | N/A |
| Divorced/Separated | N/A | Divorced/Separated | N/A |
| Widowed | N/A | Widowed | N/A |
| **Employed** |  | **Employed** |  |
| Yes | 1.00 | Yes | 1.00 |
| No | N/A | No | N/A |
| **Behaviours** |  | **Behaviours** |  |
| **Drinks alcohol** |  | **Drinks alcohol** |  |
| Yes | 1.00 | Yes | 1.00 |
| No | 0.56<br>(0.13,<br>2.39) | No | N/A |
| **ID use** |  | **ID use** |  |
| Yes | N/A | Yes | 1.00 |
| No | N/A | No | N/A |
| **Ever received HBV test** |  | **Ever received HBV test** |  |
| Yes | 1.00 | Yes | 1.00 |
| No | N/A | No | N/A |
| **Knowledge** |  | **Knowledge** |  |
| **Has heard of HBV** |  | **Has heard of HBV** |  |
| Yes | 1.00 | Yes | 1.00 |
| No | 2.78<br>(0.34,<br>22.96) | No | N/A |
| **Knows how HBV is spread** |  | **Knows how HBV is spread** |  |
| Yes | 1.00 | Yes | 1.00 |
| No | 0.72<br>(0.09,<br>6.02) | No | N/A |
| **Knowledge score** | 1.10<br>(0.93,<br>1.31) | **Knowledge score** | 2.05<br>(0.77,<br>5.45) |
| **Stigma score** | 0.79<br>(0.55,<br>1.12) | **Stigma score** | 0.80<br>(0.21,<br>2.99) |
| **Knowledge vs Stigma** | 0.99<br>(0.92,<br>1.06) | **Knowledge vs Stigma** | 1.01<br>(0.93,<br>1.09) |

When assessing the relationship between HBsAb and the variables of interest, the only variable from the Pokhara group to produce a statistically significant result was whether participants had been tested for HBV in the past (OR = 0.15 95% CI 0.04, 0.63) which indicated that participants who had not been tested were less likely to have been positive for HBsAb. In the Dolpa group, some factors which increased the likelihood of the presence of HBsAb were being older (OR = 1.05, 95% CI 1.03, 1.07) and being widowed (OR = 4.33, 95% CI 1.14, 16.49). Having a high school education (OR = 0.39, 95% CI 0.20, 0.77) decreased the likelihood of the presence of HBsAb, however this effect was not linear as this association was not shown in those with a university education. The full table of results from the HBsAb regressions are shown in Supplementary Table 2.

### Knowledge and Stigma around HBV

The majority of both samples had not heard of HBV prior to this study (72% for the Dolpa group and 57% for the Pokhara group). Despite low knowledge around HBV transmission or knowing someone who ahd been ifeceted, a relatively high level of stigma was evident in both groups. Although neither sample were well-aware of methods of transmission for HBV, there were slight differences between the responses in each area, with the Pokhara cohort choosing the “Don’t know” option more frequently than the Dolpa cohort, which chose the more definite “No” response more often. The responses for the question “Have you, or anyone you know, ever received a vaccination for HBV?” showed the vast majority of the sample believed they had not or did not know anyone who had received a vaccination, with 89% and 93% responding “No” for Dolpa and Pokhara respectively.

The questions around stigma associated with being HBV positive showed a similar pattern, with the Dolpa group being more certain in their responses and a greater proportion of the Pokhara group being less sure, or more willing to admit that they didn’t know. There were some questions which represented a major difference in perceptions of people with HBV between the two cohorts, such as the question which asked whether participants would buy food from a vendor with HBV. For the Dolpa cohort, 28% (95% CI 23.2%, 32.2%) responded “Yes”, while the 52% (95% CI 46.7%, 56.7%) of the Pokhara cohort responded that they would not have an issue with this. For a full summary of the results and questions, see Supplementary Table 4. The association between having a greater level of knowledge of HBV and the level of stigma was also assessed using linear regressions. For the Dolpa study group, there was a slight increase in stigma the more individuals knew about HBV, however the opposite was true of the Pokhara group (Supplementary Table 3).

### Healthcare service use

The responses to the questions focussing on some aspects of healthcare service use are summarised in Supplementary Table 3. The majority of both groups had not interacted with a community healthcare worker within the last 12 months (76% for Dolpa, 63% for Pokhara), however over 96% of both cohorts had received vaccinations and had visited healthcare clinics at some point in their lives. The question which represented the largest difference between the cohorts focussed on the participants’ knowledge of where to go to receive a blood test, with only 51% of the Dolpa cohort knowing where to go compared to 80% of the Pokhara cohort.

## DISCUSSION

The prevalence of HBV among the study populations was lower than anticipated, which is a positive outcome and could reflect the effectiveness of the vaccination program in Nepal in these study populations. However, the HBV prevalence was approximately 4 times higher in the more remote Dolpa area than urban Pokhara. The overall lower than expected HBV prevalence was unexpected due to the average ages of the respective cohorts, as much of the study population were already adults by the time the vaccination program was introduced in 2002-3. As children have been the focus of this program, it would have been expected that the adult population would be at a greater risk of HBV infection being present. The vast majority of each cohort believed they had not received a vaccination for HBV, although almost all had participated in vaccination programs for other infectious diseases. Additionally, most of the participants had not heard of HBV, so it is possible that they had been vaccinated for this disease without knowing specifically what it was for as the HBV vaccine is given in conjunction with other vaccines in a pentavalent vaccine (5, 8). It this therefore possible that adults who have taken their children to be vaccinated may not realise the list of diseases which are included within the pentavalent vaccines, thus potentially resulting in the scenario observed in this study

The overall knowledge level of how HBV is spread and the behaviours to minimise to avoid infection was low among study participants in both geographic catchments, consistent with the high proportion of participants who had not ever heard of HBV. However, despite this low knowledge, many participants also indicated stigma towards those who were HBV positive. It is possible that due to the relatively long-term nature of HBV infections the disease is not a priority in communities which have far more pressing concerns, especially in terms of other infectious diseases such as Covid-19 and tuberculosis. During the heights of the Covid-19 pandemic, the spreading of misinformation lead to increased levels of stigma around Covid-19, how it was spread and who was to “blame” for the spread (16, 17). Similar stigma is experienced by those with tuberculosis (18), however this stigma is not exclusive to Nepal (19). It is likely that underlying mistrust of individuals who are “sick” is the reason that although overall knowledge of HBV was low, the study population were wary of infected individuals. For Covid-19, this stigma extended even to healthcare workers and those who had recovered from infection (20).

However, there were slight differences in stigma opinions between the two cohorts. In general, the cohort based in Dolpa was more certain in their responses, opting to choose either “Yes” or “No” rather than “Don’t know”. The cohort from Pokhara appeared to be more willing to choose this option, which meant that the distribution of the answers varied between sites. The Dolpa sample showed a higher level of stigma towards those infected with HBV, with a higher proportion wishing to keep it a secret if a family member was HBV positive and not wanting to buy food from an HBV positive vendor. Within the two study populations, having tested positive for HBsAg was also associated with a higher knowledge score, which likely indicates that people are not provided information on HBV until they have been infected. Having a more positive view and a less stigmatised perception towards HBV was associated with a decreased likelihood of being positive for either HBsAg or HBsAb, although these results were all statistically insignificant. It is possible that the overall level of knowledge about HBV was potentially due to the relatively high number of illiterate individuals within the cohorts, with similar proportions having also received no formal schooling.

In terms of how each cohort accessed various healthcare services, it is notable that the majority of each of the groups had participated in government provided vaccination programs. This suggests that there is potentially good engagement in these communities for future programs, however there is a need for education around certain conditions, HBV included. Since the vaccination schedule utilises multivalent vaccines (5, 8) for several infectious diseases it is possible that individuals are not aware of all of the details for each disease included. Given the low level of knowledge of HBV within the communities, as well as the uncertainty shown in how to go about getting tested or receiving treatment within these communities (55% of the Dolpa cohort and 50% of the Pokhara cohort did not know where to receive treatment), there is a clear need for health promotion resources relating to HBV. Given the laboratory results indicate there are active cases of HBV within the community, there remains the risk of transmission facilitated by this lack of knowledge.

Although there was some level of participation in known risk factors for HBV, such as unprotected sex or getting a tattoo, the overall levels of HBV were low. It is possible that these behaviours could be explained by the age of both study populations, as other risky behaviours such as intravenous drug use was low in both groups. Combining this with the lack of knowledge around how HBV is spread and the high proportion of the study population who did not believe that they had received a vaccination for HBV, there remain clear potential avenues of HBV being spread within the community.

### Strengths and Limitations

This study has several clear strengths. The survey results indicate that while there may currently be low prevalence of active cases of HBV within the communities, overall knowledge of HBV and how it is spread is low within both sample populations. As such, there is a need to emphasise this information in future educational resources and programs to be provided to these communities. These could leverage the local women’s health community educators, as HBV is most commonly spread from mother-to-child.

There are some clear limitations with this study. Although the sample size for each cohort is appropriately large, there are some differences within the study cohorts which make them unlikely to be representative of the wider population. For example, both study populations old compared to the population median and contained a high proportion of female participants, when compared to the general population. There was an uneven distribution of males and females among the participants between the cohorts which may be due to the division of labour based on traditional gender roles. It is not necessarily clear why this occurred, although it may have been influenced by the use of the community healthcare workers who aided recruitment for the study, as they are more involved with women within the community. However the difference between male and female participants in Pokhara may have been due to social or cultural influences, as it has been reported previously that the population there are less willing to engage with field teams and researchers due to perceived issues with privacy. The average age of the participants may also be indicative of which portion of the study populations were available to participate, as the average age of the cohorts (over 35 years for Dolpa, over 47 years for Pokhara) were very high. The reported median age of Nepal is approximately 25 years, indicating that the cohorts in this study were far older than most of the population. In addition, this impacts the likelihood that the participants had received vaccinations for HBV, as the programs implemented often target children.

Additionally, due to the low level of positive cases returned, it was not possible to produce any realistic estimates for which risk factors increase the likelihood of becoming infected with HBV. This may simply be a quirk of the particular sample population, however national estimates and previous surveys of these populations suggest that there should have been more positive cases of HBV within the cohorts. It is possible that there may have been some error due to sample transport or laboratory testing procedure, however this is unlikely as this study used the same biological specimen protocols as other studies conducted by CMDN, which showed higher prevalence estimates so this is unlikely to be the cause.

## DECLARATIONS

## Ethics Approval and Consent to participate

The study was implemented in accordance with ethical approval from the Western Sydney University Humar Research Ethics Committee (WSUHREC) (H13751) and the Nepal Human Research Council (NHRC) (Proposal ID 275 2021). All participants signed written informed consent forms, and were made aware that there would be no ramifications if they declined to participate.

## Consent for publication

Not applicable.

## Availability of data and materials

The datasets used and/or analysed during the current study are available from the corresponding author on reasonable request.

## Data Availability

All data produced in the present study are available upon reasonable request to the authors.

## Acknowledgement

Not applicable.

## Authors’ contributions

SH completed the ethics applications, contributed to project conception, designed the questionnaires, performed all data analysis and was a major contributor the manuscript.

KM aided in WSU ethics process, provided major contributions to project conception and final manuscript.

SD provided major contributions to project conception, in-country logistics and Nepal ethics process, and helped facilitate laboratory procedures, as well as contributing to the final manuscript.

AP aided in WSU ethics process, provided major contributions to project conception and final manuscript.

