## Supplemental table 3 for "Knowledge, Stigma and Prevalence of HBV among two populations from Nepal: a cross-sectional study"

**Supplementary Table 3.** Responses to the knowledge and stigma-focussed questions within the questionnaire.

| Stigma |  | Dolpa |  |  | Pokhara | | | | | |
| --- | --- | --- | --- | --- | --- | --- | --- | --- | --- | --- |
|  |  | **n (%)** |  |  | **n (%)** | | | | | |
|  |  | Yes | No | Don't know | Yes | | No | | Don't know | |
| If you knew a family member of yours had HBV, would you be willing to have them in your household? |  | 263 (66%) | 85 (21%) | 51 (13%) | 264 (66%) | | 33 (8%) | | 102 (26%) | |
| If a family member of yours had HBV, would you want to keep it a secret? |  | 210 (53%) | 143 (36%) | 46 (12%) | 185 (46%) | | 119 (30%) | | 96 (24%) | |
| If a shopkeeper/food-seller had HBV, would you be willing to buy food from them? |  | 110 (28%) | 226 (57%) | 62 (16%) | 207 (52%) | | 61 (15%) | | 132 (33%) | |
| Do you think people with HBV should be allowed to continue working in jobs where they may get cut? |  | 118 (30%) | 218 (55%) | 62 (16%) | 201 (50%) | | 52 (13%) | | 147 (37%) | |
| Do you think children with HBV should be able to go to the same schools as those who don't? |  | 170 (43%) | 147 (37%) | 82 (21%) | 230 (58%) | | 44 (11%) | | 126 (31%) | |
| Knowledge |  |  |  |  |  | |  | |  | |
| Have you ever heard of HBV? |  | 112 (28%) | 287 (72%) | N/A | 173 (43%) | | 227 (57%) | | N/A | |
| Have you, or someone you know, ever received a vaccination for HBV? |  | 45 (11%) | 353 (89%) | N/A | 29 (7%) | | 371 (93%) | | N/A | |
| Do you know how HBV is spread? |  | 37 (9%) | 359 (91%) | N/A | 66 (17%) | | 331 (83%) | | 3 (<1%) | |
| Can HBV be spread from contact with bodily fluids? |  | 65 (16%) | 165 (41%) | 169 (43%) | 63 (16%) | 113 (28%) | | 223 (56%) | | |
| Can HBV be spread from mother-to-child during birth and during delivery? |  | 62 (15%) | 126 (32%) | 211 (53%) | 69 (17%) | 72 (18%) | | 259 (65%) | | |
| Can HBV be spread from mother-to-child during breastfeeding? |  | 112 (28%) | 154 (39%) | 133 (33%) | 89 (22%) | 79 (20%) | | 232 (58%) | | |
| Are children more likely to be infected than adults? |  | 87 (22%) | 170 (43%) | 141 (35%) | 92 (23%) | 79 (20%) | | 229 (57%) | | |
| Can a person protect themselves from HBV by using a condom correctly during sexual intercourse? |  | 62 (15%) | 177 (45%) | 157 (40%) | 51 (25%) | 77 (19%) | | 272 (68%) | | |
| Is there a cure for HBV? |  | 59 (15%) | 180 (45%) | 160 (40%) | 205 (51%) | 45 (11%) | | 150 (38%) | | |
| Can you get HBV from mosquito bites? |  | 65 (17%) | 153 (39%) | 173 (44%) | 117 (29%) | 83 (21%) | | 200 (50%) | | |
| Do you think that a healthy-looking person could be infected with HBV? |  | 64 (16%) | 166 (42%) | 166 (42%) | 125 (31%) | 90 (23%) | | 185 (46%) | | |
| Can HBV be spread through a blood transfusion? |  | 66 (17%) | 164 (42%) | 163 (41%) | 123 (31%) | 59 (15%) | | | | 217 (54%) |
| Is it possible for someone in your community to have a confidential HBV test? |  | 40 (10%) | 198 (50%) | 160 (40%) | 71 (18%) | 152 (38%) | | | | 177 (44%) |

**Supplementary Table 4.** Responses to the healthcare utilisation questions.

| Healthcare engagement | Dolpa |  |  |  | Pokhara |  |  |
| --- | --- | --- | --- | --- | --- | --- | --- |
|  | **n (%)** |  |  |  | **n (%)** |  |  |
|  | Yes | No | Don't know | | Yes | No | Don't know |
| Have you ever participated in any vaccination programs? | 392 (98%) | 6 (2%) | 1 (<1%) |  | 383 (96%) | 15 (4%) | 2 (<1%) |
| Have you ever visited a healthcare centre/clinic? | 389 (97%) | 9 (3%) | N/A |  | 383 (96%) | 17 (4%) | N/A |
| Have you met, discussed, or interacted with any PE/OE/CM/CE in the last 12 months? | 23 (6%) | 303 (76%) | 70 (18%) |  | 23 (6%) | 253 (63%) | 124 (31%) |
| Have you ever had a blood test? | 256 (64%) | 143 (36%) | N/A |  | 330 (83%) | 69 (17%) | 1 (<1%) |
| Have you ever heard of the prevention of mother-to-child transmission services (PMTCT) for pregnant women? | 10 (3%) | 389 (97%) | N/A |  | 44 (11%) | 356 (89%) | N/A |
| Do you know where to go to received a blood test? | 202 (51%) | 197 (49%) | N/A |  | 319 (80%) | 81 (20%) | N/A |
| Do you know where to go to receive medical treatment for HBV? | 178 (45%) | 220 (55%) | N/A |  | 197 (49%) | 201 (50%) | 2 (<1%) |
