## Supplemental table 2 for "Knowledge, Stigma and Prevalence of HBV among two populations from Nepal: a cross-sectional study"

**Supplementary Table 2.** Odds ratios from univariate regression models examining the association between various demographic, behavioural and social risk factors and the likelihood of having HBsAb present within an individual.

| Dolpa HBsAb ORs | |  |  |  | Pokhara HBsAb ORs | |  |
| --- | --- | --- | --- | --- | --- | --- | --- |
| Demographics | | **Odds Ratio (95% CI)** | |  | **Demographics** | | **Odds Ratio (95% CI)** |
| Age |  | **1.05 (1.03, 2.05)** | |  | Age |  | 0.99 (0.96, 1.02) |
| Sex |  |  |  |  | Sex |  |  |
|  | Female | 1.00 |  |  |  | Female | 1.00 |
|  | Male | 1.72 (0.98, 3.01) | |  |  | Male | 2.04 (0.85, 4.98) |
| Literacy |  |  |  |  | Literacy |  |  |
|  | Illiterate | 1.00 |  |  |  | Illiterate | 1.00 |
|  | Literate | 0.60 (0.34, 1.08) | |  |  | Literate | 1.54 (0.55, 4.31) |
| Education level | |  |  |  | Education level | |  |
|  | No formal schooling | 1.00 |  |  |  | No formal schooling | 1.00 |
|  | Primary | 0.93 (0.44, 1.93) | |  |  | Primary | 0.78 (0.22, 2.78) |
|  | High school | **0.39 (0.20, 0.77)** | |  |  | High school | 2.04 (0.64, 6.47) |
|  | University | 1.01 (0.31, 3.31) | |  |  | University | 7.20 (1.49, 34.78) |
| Marital Status | |  |  |  | Marital Status | |  |
|  | Never married | 1.00 |  |  |  | Never married | 1.00 |
|  | Married | 1.05 (0.39, 2.83) | |  |  | Married | 0.32 (0.10, 1.03) |
|  | Divorced/Separated | 1.70 (0.16, 18.58) | |  |  | Divorced/Separated | N/A |
|  | Widowed | **4.33 (1.14, 16.49)** | |  |  | Widowed | N/A |
| Employed |  |  |  |  | Employed |  |  |
|  | Yes | 1.00 |  |  |  | Yes | 1.00 |
|  | No | 1.14 (0.51, 2.55) | |  |  | No | 2.92 (0.86, 7.34) |
| Behaviours | |  |  |  | **Behaviours** | |  |
| Drinks alchohol | |  |  |  | Drinks alchohol | |  |
|  | Yes | 1.00 |  |  |  | Yes | 1.00 |
|  | No | 0.71 (0.39, 1.32) | |  |  | No | 1.37 (0.39, 4.79) |
| ID use |  |  |  |  | ID use |  |  |
|  | Yes | 1.00 |  |  |  | Yes | 1.00 |
|  | No | N/A |  |  |  | No | N/A |
| Ever received HBV test | | |  |  | Ever received HBV test | | |
|  | Yes | 1.00 |  |  |  | Yes | 1.00 |
|  | No | N/A |  |  |  | No | **0.15 (0.04, 0.63)** |
| Knowledge | |  |  |  | **Knowledge** | |  |
| Has heard of HBV | |  |  |  | Has heard of HBV | |  |
|  | Yes | 1.00 |  |  |  | Yes | 1.00 |
|  | No | 1.03 (0.55, 1.92) | |  |  | No | 0.83 (0.34, 2.01) |
| Knows how HBV is spread | | |  |  | Knows how HBV is spread | | |
|  | Yes | 1.00 |  |  |  | Yes | 1.00 |
|  | No | 1.46 (0.50, 4.30) | |  |  | No | 1.21 (0.34, 4.24) |
| Knowledge score | | 0.94 (0.85, 1.03) | |  | Knowledge score | | 2.05 (0.77, 5.45) |
| Stigma score | | 0.99 (0.87, 1.12) | |  | Stigma score | | 0.80 (0.21, 2.99) |
