## Supplemental table 1 for "Knowledge, Stigma and Prevalence of HBV among two populations from Nepal: a cross-sectional study"

**Supplementary Table 1.** The full results of the blood tests are shown here.

| HBV Results table | |  |  |  |
| --- | --- | --- | --- | --- |
|  |  | **Dolpa** |  | **Pokhara** |
| HBsAg | Positive | 8 |  | 2 |
|  | Negative | 391 |  | 397 |
| HBsAb | Positive | 58 |  | 21 |
|  | Negative | 341 |  | 379 |
| HBeAg | Positive | 3 |  | 3 |
|  | Negative | 394 |  | 397 |
| HBeAb | Positive | 4 |  | 3 |
|  | Negative | 395 |  | 396 |
| HBcAb | Positive | 10 |  | 3 |
|  | Negative | 388 |  | 394 |
